# THE AIRBORNE CONTAGIOUSNESS OF RESPIRATORY VIRUSES: A COMPARATIVE ANALYSIS AND IMPLICATIONS FOR MITIGATION

**DOI:** 10.1101/2021.01.26.21250580

**Authors:** A. Mikszewski, L. Stabile, G. Buonanno, L. Morawska

**Author notes:** Corresponding Author: Professor Lidia Morawska, International Laboratory for Air Quality and Health, Queensland University of Technology, 2 George Street, Brisbane, Queensland 4001, Australia.

## Abstract

**Background:** The infectious emission rate is a critical input parameter for airborne contagion models, but data are limited due to reliance on estimates from chance superspreading events. A predictive estimation approach for the quanta emission rate (ER_q_) was recently proposed for SARS-CoV-2 using the droplet volume concentration of various expiratory activities. This study assesses the strength of the approach and uses novel predictive estimates of ER_q_ to compare the contagiousness of respiratory pathogens.

**Methods:** We applied the predictive approach to SARS-CoV-1, SARS-CoV-2, MERS, measles virus, adenovirus, rhinovirus, coxsackievirus, seasonal influenza virus and *Mycobacterium tuberculosis* (TB) and compared ER_q_ estimates to values reported in literature. We calculated infection risk in a prototypical classroom and barracks to assess the relative ability of ventilation to mitigate airborne transmission.

**Results:** Our median standing and speaking ER_q_ estimate for SARS-CoV-2 (2.6 quanta hour (h)^-1^) is similar to active, untreated TB (3.1 h^-1^), higher than seasonal influenza (0.17 quanta h^-1^), and lower than measles virus (15 quanta h^-1^). We calculated event reproduction numbers above 1 for SARS-CoV-2, measles virus, and untreated TB in both the classroom and barracks for an activity level of standing and speaking at low, medium and high ventilation rates of 2.3, 6.6 and 14 liters per second per person, respectively.

**Conclusions:** Our predictive ER_q_ estimates are consistent with the range of values reported over decades of research. In congregate settings, current ventilation standards are unlikely to control the spread of viruses with upper quartile ER_q_ values above 10 quanta h^-1^, such as SARS-CoV-2, indicating the need for additional control measures.

## 1.0 Introduction

The COVID-19 pandemic has renewed attention to airborne contagion in shared indoor atmospheres. Airborne transmission of respiratory tract infection results from the inhalation of virus- or bacteria-laden droplet nuclei, defined as the evaporated residua of respiratory droplets expired during breathing, vocalizing, coughing, and sneezing. Modeling by Balachandar et al. [1] indicates that all expired respiratory droplets below 100 μm in diameter will evaporate to droplet nuclei of non-volatile biological material within a second of expiration and after less than 1 meter of travel, even at 98% ambient air humidity. As such, there is an urgent need to quantify the emission rate of droplets below 100 μm to facilitate airborne infection risk assessment. Buonanno et al. [2] developed a novel predictive estimation approach for the quanta emission rate as a function of respiratory activity and activity level. The quantal dose-response concept for airborne contagion was originally developed by Wells [3] with the understanding that infection by inhalation is a probabilistic process involving myriad random variables with substantial heterogeneity. Using a Poisson model, a quantum equals the unknown *amount* of pathogenic airborne droplet nuclei that will cause sustained infection in 63% of exposed susceptibles. As cautioned by Nardell [4], “quantum,” the dose traditionally back calculated from the end result of infections in a group of susceptibles, should not be confused with “infectious particles,” that originate from the infected source and can be measured in units of RNA copies or plaque forming units (PFUs). The predictive estimation approach bridges the gap between these two concepts, with a quantum representing a human infectious dose for 63% of susceptibles (HID_63_) by droplet nuclei inhalation that can be approximately related to a viral or bacillary load in the emitting subject through experimental analysis, as was achieved for seasonal influenza virus by Bueno de Mesquita et al. [5].

The aim of this work is threefold: 1) to assess the strength of the predictive estimation approach for the airborne emission rate of common respiratory pathogens by comparing estimates to back-calculated values reported in literature, 2) to use the estimates to compare the contagiousness of the modeled pathogens through the airborne route, and 3) to assess the ability of modern standards of ventilation to prevent their epidemic spread.

## 2. Materials and methods

The predictive estimation approach for the quanta emission rate (ER_q_) is presented as equation (1) for respiratory viruses [6]:

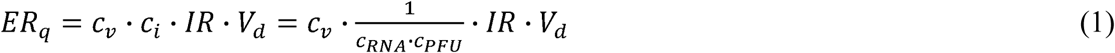

where c_v_ is the viral load in sputum (RNA copies mL^-1^), c_i_ (quanta RNA copies^-1^) is a conversion factor defined as the ratio between one quantum and the infectious dose expressed in viral RNA copies, *IR* is the inhalation rate (m^3^ h^-1^), and *V*_*d*_ is the droplet volume concentration expelled by the infectious person (mL m^-3^). The conversion factor, c_i_, can be calculated as the product of the number of RNA copies per plaque-forming unit (PFU) (c_RNA_) and the number of PFU approximating the human infective dose (HID_63_) by droplet nuclei inhalation (one quantum) (c_PFU_, PFU quanta^-1^). Where viral load is provided in units of PFU mL^-1^, or the median tissue culture infectious dose (TCID_50_) mL^-1^, the C _RNA_ term becomes unnecessary. For unit conversions, approximately four-fifths of a quantum is a TCID_50_ [3], and a PFU is commonly approximated as seven-tenths of a TCID_50_. The droplet volume concentration *V*_*d*_ is a function of the expiratory activities and was derived from the total volume emitted by a loud-speaking person provided by Stadnytskyi et al. [7]. Representative values for the inhalation rate (0.49 m^3^ h^-1^ for resting, 0.54 m^3^ h^-1^ for standing, and 1.38 m^3^ h^-1^ for light exercise) were obtained from Adams [8].

While inhalation rate and droplet volume concentrations are known to vary between individuals based on age, body mass, and natural physiological heterogeneity, we hold them constant for each of three evaluated expiratory activities to simplify the calculation and limit variation to the viral load. With this assumption the product *IR* · *V*_*d*_ becomes a constant droplet volume emission rate in mL h^-1^ as follows: 9.8 x 10^−4^ for resting, oral breathing; 4.9 x 10^−3^ for standing, speaking; and 8.3 x 10^−2^ for light activity, speaking loudly. Derived from a laser light scattering study [7], the droplet emission rates span the size range of expiratory droplets produced by speaking. The high sensitivity of light scattering better quantifies droplets in the range of 10 to 100 μm which can be missed by aerodynamic particle sizer (APS) measurements.

To apply the predictive approach, we used equation (1) and a lognormal distribution of viral load to create a lognormal distribution of ER_q_ defined by an associated probability density function. We compiled viral load and infectious dose data for the following eight (8) viruses: severe acute respiratory syndrome (SARS) coronavirus (CoV) 1 and 2, Middle East respiratory syndrome (MERS) coronavirus, measles virus, seasonal influenza virus, rhinovirus, coxsackievirus, and adenovirus. We did not include respiratory syncytial virus (RSV) in our evaluation because transmission between infants is generally a greater concern than between adults and this is beyond our current scope [9]. We did not include smallpox (variola) virus, chickenpox (varicella) virus, parainfluenza viruses, mumps and rubella viruses and others due to lack of data on viral load in the respiratory tract and/or infectivity by droplet nuclei inhalation. While the focus of this paper is respiratory viruses, we did apply the predictive approach to *Mycobacterium tuberculosis* (TB) as it is the most well-studied agent of airborne contagion. For TB we separately used bacillary loads representative of both untreated active cases and cases after two weeks of treatment, and an estimate of infectious dose in colony forming units (CFUs). Parameters selected for each virus and for TB are summarized in Table 1. A narrative with referencing for all parameter values and rationales for inclusion is provided in the Supplemental Material. Where sputum viral load data were unavailable or considered otherwise non-representative, we used nasal swab or nasal wash data as a substitute.

**Table 1.** Viral/bacillary load and infectivity input data

| Pathogen | $\log_{10} c_v$<br>average (st.dev) | Conversion Factor ($c_i$) |
| --- | --- | --- |
| Adenovirus | 3.2 (0.95) TCID<sub>50</sub> mL<sup>-1</sup> | 0.50 quanta TCID<sub>50</sub><sup>-1</sup> |
| Coxsackievirus | 3.4 (1.1) TCID<sub>50</sub> mL<sup>-1</sup> | 0.025 quanta TCID<sub>50</sub><sup>-1</sup> |
| Influenza | 6.7 (0.84) RNA copies mL<sup>-1</sup> | 7.1 x 10<sup>-6</sup> quanta RNA copies<sup>-1</sup> |
| Measles | 3.5 (1.6) TCID<sub>50</sub> mL<sup>-1</sup> | 1.0 quanta TCID<sub>50</sub><sup>-1</sup> |
| MERS | 6.7 (1.6) RNA copies mL<sup>-1</sup> | 2.3 x 10<sup>-6</sup> quanta RNA copies<sup>-1</sup> |
| Rhinovirus | 3.6 (0.83) TCID<sub>50</sub> mL<sup>-1</sup> | 0.053 quanta TCID<sub>50</sub><sup>-1</sup> |
| SARS-CoV-1 | 6.1 (1.3) RNA copies mL<sup>-1</sup> | 6.8 x 10<sup>-6</sup> quanta RNA copies<sup>-1</sup> |
| SARS-CoV-2 | 5.6 (1.2) RNA copies mL<sup>-1</sup> | 1.4 x 10<sup>-3</sup> quanta RNA copies<sup>-1</sup> |
| TB (Untreated) | 5.5 (1.3) CFU mL<sup>-1</sup> | 2.0 x 10<sup>-3</sup> quanta CFU<sup>-1</sup> |
| TB (On Treatment) | 4.0 (1.4) CFU mL<sup>-1</sup> |  |

At the time of writing this paper, there is mounting concern regarding the emergence of novel strains of SARS-CoV-2 that preliminary epidemiological data suggest are more transmissible than those circulating previously. A more contagious strain would have higher ER_q_ values through a higher median viral load and/or a lower infectious dose. Our calculations used the thermodynamic equilibrium dose-response model of Gale [10], which suggests a very low median infectious dose of only 1 to 2 PFU due to the relative absence of a protective effect in the mucus barrier. Our ER_q_ estimates already reflect the possibility of high contagiousness of SARS-CoV-2 and we do not expect forthcoming data on different strains to significantly change the conclusions presented herein.

To quantify the ability of ventilation to mitigate the risk of airborne transmission, we calculated the individual risk (R) of an exposed person [6] and event reproduction numbers (R_event_) for a typical classroom studied by Wells [11] and for a military barracks studied by Couch et al. [12]. R_event_ is defined as the expected number of new infections arising from a single infectious individual at an event [13]. We performed the calculations using low, medium, and high ventilation rates of 2.3 liters per second per person (L s^-1^ p^-1^), 6.6 L s^-1^ p^-1^, and 14 L s^-1^ p^-1^, respectively, and assuming one infectious occupant and a fully susceptible population. Our low and medium ventilation rates correspond to the average values estimated for low and high ventilation dormitories in Zhu et al. [14], and our high ventilation rate corresponds to the value estimated by Wells for the control classrooms in his air disinfection experiments [11]. For reference, the ANSI/ASHRAE 62.1 combined outdoor air rate values for acceptable indoor air quality for these two spaces are approximately 6.3 L s^-1^ p^-1^ and 3.7 L s^-1^ p^-1^ for classrooms and barracks sleeping areas, respectively [15]. A summary of the modeling approach and parameter assumptions are provided in the Supplemental Material.

## 3. Results & Discussion

### 3.1 Quanta Emission Rates

The median ER_q_ estimates from the predictive estimation approach are ranked from high to low as follows: measles virus, adenovirus, TB (untreated), SARS-CoV-2, rhinovirus, coxsackievirus, seasonal influenza, TB (on treatment), MERS, and SARS-CoV-1. Table 2 provides the 10^th^ percentile, 50^th^ percentile (median), and 90^th^ percentile ER_q_ estimates for each virus for the three emission profiles evaluated herein (resting, oral breathing; standing, speaking; light activity, speaking loudly). Based on the assumed lognormal distribution for the viral load, the ER_q_ estimates also follow a lognormal distribution, with the log_10_ average value equal to the log_10_ of the reported median value in Table 2, and the log_10_ standard deviation equal to that of the viral load in Table 1. For example, the log_10_ ER_q_ distribution for SARS-CoV-2 for standing and speaking has an average of 0.41 (the log_10_ of 2.6) and a standard deviation of 1.2. Plots of the lognormal distributions for the standing and speaking activity level are provided in Figure 1. Published ER_q_ values in literature are presented in Table 3 for comparison.

**Table 2.**
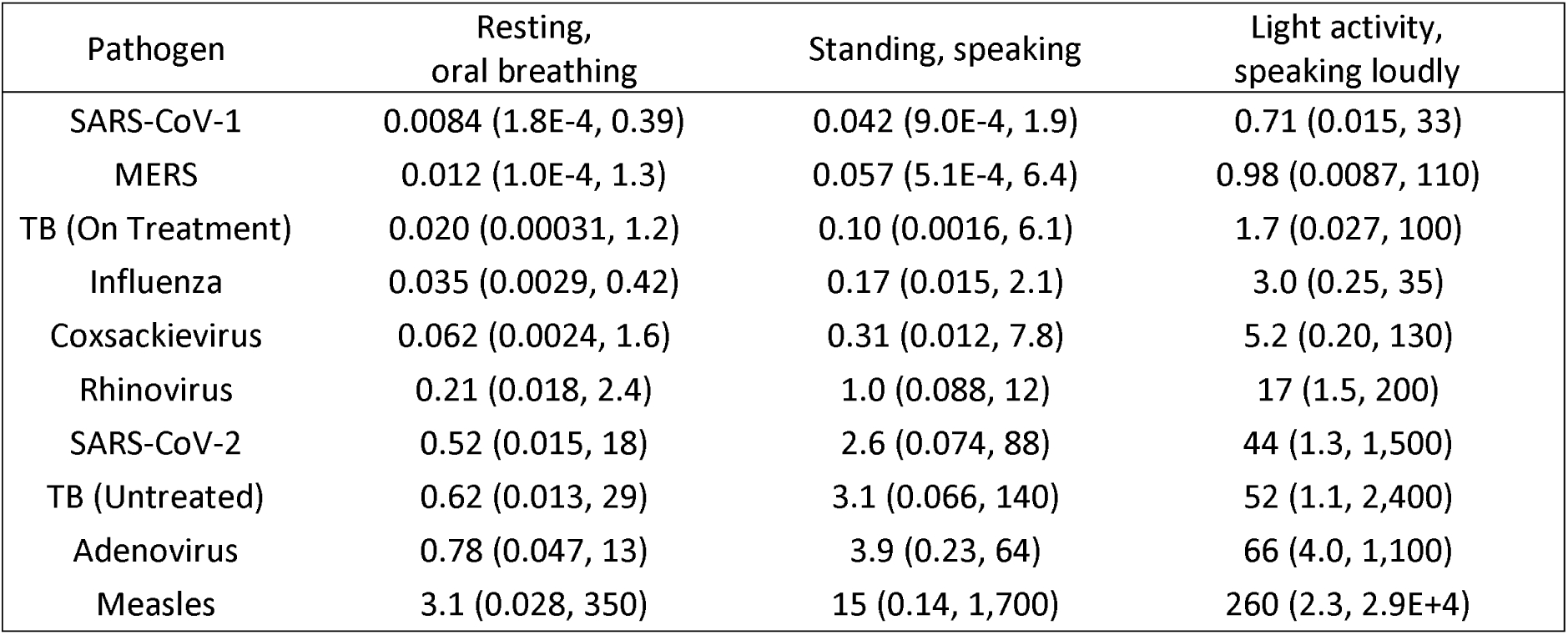
Predictive ER _q_(quanta h^-1^) statistics as a function of the expiratory activity and activity level. *50^th^ percentile (10^th^ percentile, 90^th^ percentile)*

| Pathogen | Resting,<br>oral breathing | Standing, speaking | Light activity,<br>speaking loudly |
| --- | --- | --- | --- |
| SARS-CoV-1 | 0.0084 (1.8E-4, 0.39) | 0.042 (9.0E-4, 1.9) | 0.71 (0.015, 33) |
| MERS | 0.012 (1.0E-4, 1.3) | 0.057 (5.1E-4, 6.4) | 0.98 (0.0087, 110) |
| TB (On Treatment) | 0.020 (0.00031, 1.2) | 0.10 (0.0016, 6.1) | 1.7 (0.027, 100) |
| Influenza | 0.035 (0.0029, 0.42) | 0.17 (0.015, 2.1) | 3.0 (0.25, 35) |
| Coxsackievirus | 0.062 (0.0024, 1.6) | 0.31 (0.012, 7.8) | 5.2 (0.20, 130) |
| Rhinovirus | 0.21 (0.018, 2.4) | 1.0 (0.088, 12) | 17 (1.5, 200) |
| SARS-CoV-2 | 0.52 (0.015, 18) | 2.6 (0.074, 88) | 44 (1.3, 1,500) |
| TB (Untreated) | 0.62 (0.013, 29) | 3.1 (0.066, 140) | 52 (1.1, 2,400) |
| Adenovirus | 0.78 (0.047, 13) | 3.9 (0.23, 64) | 66 (4.0, 1,100) |
| Measles | 3.1 (0.028, 350) | 15 (0.14, 1,700) | 260 (2.3, 2.9E+4) |

**Table 3.** Predictive ER_q_ comparisons with literature values

| Virus | Setting | ER<sub>q</sub><br>(quanta h<sup>-1</sup>) | Standing,<br>speaking<br>(percentile) | Light activity,<br>speaking loudly<br>(percentile) |
| --- | --- | --- | --- | --- |
| SARS-CoV-1 | Outbreak in Taipei Hospital [16] | 29 | 98<sup>th</sup> | 89<sup>th</sup> |
| SARS-CoV-2 | Wuhan Apartment [17] | 15 | 74<sup>th</sup> | 35<sup>th</sup> |
|  | Cruise Ship [17] | 15 | 74<sup>th</sup> | 35<sup>th</sup> |
|  | Wuhan Bus #1 [18] | 36 | 83<sup>rd</sup> | 47<sup>th</sup> |
|  | Restaurant [6] | 61 | 87<sup>th</sup> | 55<sup>th</sup> |
|  | Wuhan Bus #2 [18] | 62 | 88<sup>th</sup> | 55<sup>th</sup> |
|  | School, Germany [19] | 116 | 92<sup>nd</sup> | 64<sup>th</sup> |
|  | Buddhist Bus [18] | 133 | 92<sup>nd</sup> | 66<sup>th</sup> |
|  | School, Israel [19] | 139 | 92<sup>nd</sup> | 66<sup>th</sup> |
|  | Meeting [19] | 139 | 92<sup>nd</sup> | 66<sup>th</sup> |
|  | Fitness Center [18] | 152 | 93<sup>rd</sup> | 67<sup>th</sup> |
|  | Abattoir [19] | 232 | 95<sup>th</sup> | 73<sup>rd</sup> |
|  | Call Center [18] | 683 | 98<sup>th</sup> | 84<sup>th</sup> |
|  | Chorus, USA [20] | 970 | 98<sup>th</sup> | 87<sup>th</sup> |
|  | Chorus, Germany [19] | 4,213 | -- | 95<sup>th</sup> |
| Measles | Classroom – average child with measles [3, 21] | 18 | 52<sup>nd</sup> | 23<sup>rd</sup> |
|  | Retrospective analysis of elementary and secondary school outbreaks [22] | 60 (minimum) | 64<sup>th</sup> | 34<sup>th</sup> |
|  |  | 600 (median) | 84<sup>th</sup> | 59<sup>th</sup> |
|  |  | 5,600 (maximum) | 94<sup>th</sup> | 80<sup>th</sup> |
|  | Outbreak in a secondary school [23] | 2,765 | 92<sup>nd</sup> | 74<sup>th</sup> |
|  | Outbreak in pediatrician's office [24] | 8,640 | 96<sup>th</sup> | 83<sup>rd</sup> |
| Influenza | Human transmission trials in quarantine rooms [5] | 0.11 | 40<sup>th</sup> | 5<sup>th</sup> |
|  | Transmission experiments among ferrets [25] | 7.95 | 97<sup>th</sup> | 70<sup>th</sup> |
|  | Airliner outbreak during delay with inoperable ventilation [26, 27] | 79 | -- | 95<sup>th</sup> |
| Rhinovirus | Transmission trials using card playing games [26, 28] | 2.0 | 64<sup>th</sup> | 13<sup>th</sup> |

**Figure 1.**
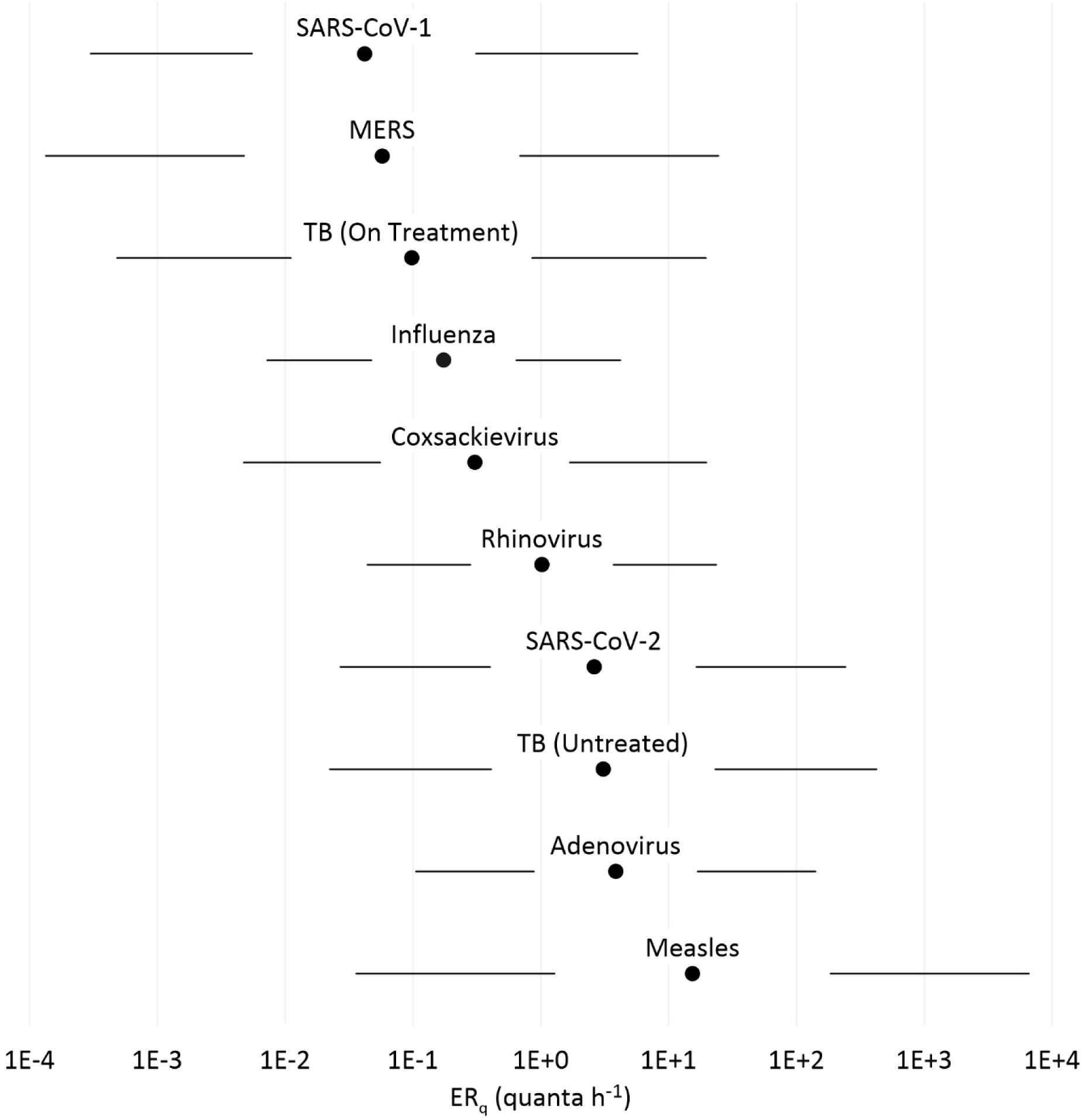
Predictive ER_q_ distributions for the standing and speaking activity level. Labeled dots are the median estimates with lines to the left of the labels spanning the 5^th^-25^th^ percentile ranges and lines to the right of the labels spanning the 75^th^-95^th^ percentile ranges.

The three evaluated coronaviruses have similar viral load distributions, but the significant difference in infectivity results in ER_q_ estimates for SARS-CoV-2 that are over an order of magnitude higher than SARS-CoV-1 and MERS. Recent estimates of ER_q_ back calculated from SARS-CoV-2 superspreading events are presented in Table 3, including data from pre-prints that are subject to revision, and approximately span the 40^th^-90^th^ percentile for the light activity, speaking loudly distribution.

Previous ER_q_ estimates for measles virus are significantly different based on calculations made before and after the introduction of the vaccine in the early 1960s. Riley et al. [21] reported the emission rate of the average child with measles to be 18 quanta h^-1^ based on the earlier work of Wells [3, 11], which is very similar to our median estimate for standing and speaking. The pattern of the spread of measles in schools studied by Wells in the pre-vaccine era was consistent with decades of epidemiology showing outbreaks to begin when the density of susceptibles reached 30-40%, and wane when the density decreased to 15-20% [3, 11]. Conversely, the post-vaccine era estimates [22-24] are based on superspreading events with back calculated emission rates reaching over 1,000 quanta h^-1^. This discrepancy is potentially explained by the impact of the density of susceptibles on the threshold emission rate needed to reproduce infection. Wells [3] noted a contact rate, or probability of infection, of 11% for measles over a three-day infectious period in a well-ventilated classroom, from which the 18 quanta h^-1^ estimate was derived. The initial density of susceptibles in the class was approximately 33%. If this density of susceptibles were reduced to 5% through vaccination, to generate an equivalent number of secondary cases on average, the contact rate would need to increase to approximately 75%. This corresponds to an emission rate over 200 quanta h^-1^, more consistent with the post-vaccine estimates of Riley [22]. Community transmission of measles in the post-vaccine era appears limited to individuals with high viral load, and thus high ER_q_, capable of picking out the few remaining susceptibles in a group [29].

The median estimated ER_q_ for seasonal influenza for standing, speaking (0.17 quanta h^-1^) is consistent with the recent estimate of 0.11 quanta h^-1^ calculated from a human transmission trial [5]. The 79 quanta h^-1^ estimate for a superspreading event on a grounded airliner [26, 27] is equal to the 95^th^ percentile value for the light activity, speaking loudly activity level (Table 3). Supporting the extreme nature of the airliner case study, Bischoff et al. [30] measured the maximum emission rate from 61 influenza patients to be approximately 1.2 x 10^6^ RNA copies h^-1^, equal to 8.7 quanta h^-1^ using the conversion factor in Table 1. We therefore conclude that ER_q_ values above 10 quanta h^-1^ may be quite uncommon for seasonal influenza, limiting the potential for explosive outbreaks of short duration. However, our estimates are for breathing and vocalizing, and severely symptomatic cases with high frequency of cough may generate significantly higher emissions, as with the airliner case study. Similarly, our estimates do not apply to a pandemic influenza with much higher susceptibility in the population.

The median estimated ER_q_ for rhinovirus of 1.0 quanta h^-1^ for standing and speaking is consistent with the geometric mean value of 2.0 quanta h^-1^ calculated based on the range of values (0.6 – 7.8) estimated by Rudnick and Milton [26].

The median ER_q_ estimated for adenovirus of 3.9 quanta h^-1^ for standing and speaking is second only to measles. No literature values are available for comparison, but a high emission rate is consistent with explosive outbreaks observed at US military basis during the late 1990s and early 2000s when the adenovirus vaccine was temporarily unavailable. Russell et al. [31] describe one such military outbreak where over the course of a 4-week period a 98% attack rate was observed among 180 susceptible persons. Echavarria et al. [32] identified a correlation between adenovirus PCR results on air filters and the number of hospitalizations within military companies and found that companies with one ventilation unit per floor and wing had lower attack rates (11%) than those with one ventilator supplying air for multiple floors (18-21%). While resumption of adenovirus vaccination largely eliminated these types of outbreaks on US military bases, China has not yet included adenovirus in its military vaccination program. Guo et al. [33] described a recent outbreak of adenovirus type 7 at a boot camp in China that lasted 30 days, resulting in 375 cases and 109 hospitalizations.

The estimated ER_q_ values for coxsackievirus fall between those of influenza and rhinovirus, with a median of 0.31 quanta h^-1^ for standing and speaking. Airborne transmission of coxsackievirus A_21_ was conclusively demonstrated in a cross-infection experiment conducted in a military barracks with physical separation of occupants and well-mixed air [12]. We modeled the average emission rate for each of 10 infected subjects in this experiment to be between 1.3 and 3.6 quanta h^-1^, consistent with the predictive calculations at the 72^nd^ to 83^rd^ percentile range of the standing and speaking distribution. A description of the experiment and our modeling approach is provided in the Supplemental Material.

The closest model for the contagiousness of SARS-CoV-2 may be provided by an untreated, active case of TB, as we estimated a median ER_q_ of 3.1 quanta h^-1^ for standing and speaking, approximately 20% higher than that of SARS-CoV-2. The office outbreak from an untreated case modeled by Nardell et al. [34] (13 quanta h^-1^) corresponds to the 68^th^ percentile of the standing, speaking distribution, while the modeled emission rate from an explosive outbreak of multi-drug resistant (MDR) TB aboard a long-haul flight (108 quanta h^-1^ [35]) corresponds to the 88^th^ percentile value. A comparison of the standing and speaking ERq distributions for TB and SARS-CoV-2 is presented in Figure 2, along with TB ER_q_ estimates from the original human-to-guinea pig transmission trials of Riley et al. [21, 36] and the similar studies in the MDR-TB and HIV era [37, 38].

**Figure 2.**
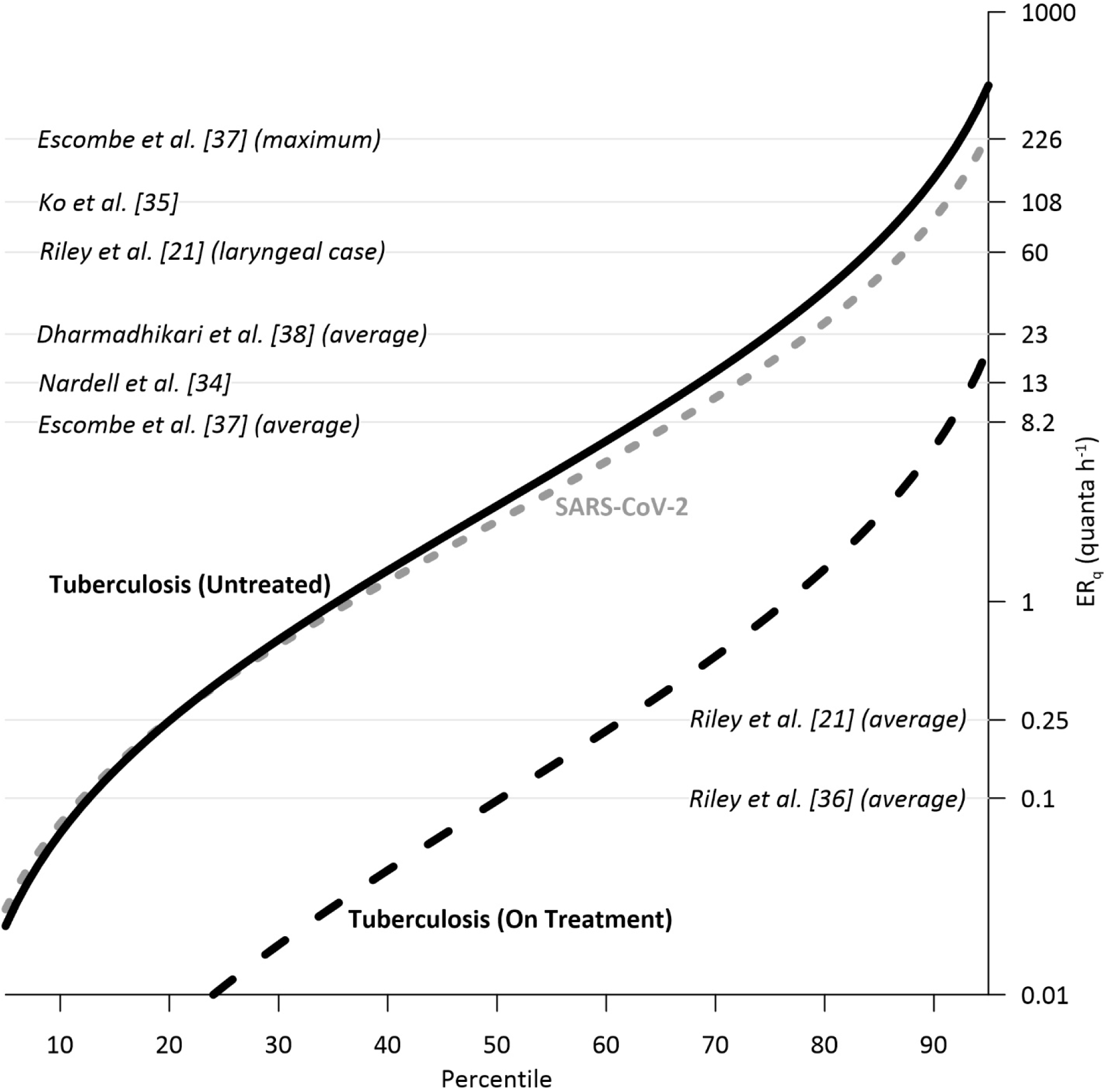
Predictive ER_q_ distributions for tuberculosis (TB) and SARS-CoV-2 for standing and speaking as compared to literature values for TB published between 1959 and 2012.

### 3.2 Classroom and Barracks Modeling Scenarios

Results of the two modeling scenarios described in the Supplemental Material are presented in Figure 3a (classroom) and Figure 3b (barracks). With respect to the individual risk (R) in both settings at the standing and speaking expiration, the results indicate: 1) the low ventilation rate (2.3 L s^-1^ p^-1^) is only able to keep R below 1% for seasonal influenza and SARS-CoV-1, 2) the high ventilation rate (14 L s^-1^ p^-1^) is needed to keep R approximately at or below 1% for rhinovirus, MERS, coxsackievirus, and TB (on treatment), and 3) even at the high ventilation rate, R is above 1% for adenovirus, TB (untreated), and SARS-CoV-2, and above 10% for measles. With respect to the expected number of infections resulting from the exposures at standing and speaking: 1) at the high ventilation rate, R_event_ is above 1 in both settings for TB (untreated), SARS-CoV-2, and measles, with adenovirus also above 1 in the barracks, and 2) R_event_ approaches or exceeds 1 in the barracks at the low ventilation rate for rhinovirus, MERS, coxsackievirus, and TB (on treatment). Illustrating the potential for high attack rates of SARS-CoV-2 in congregate housing, R_event_ is above 1 in the barracks at the resting, oral breathing expiration at both the medium and low ventilation rates. The four pathogens with calculated R_event_ values above 1 at the high ventilation rate have upper quartile ER_q_ estimates above 10 quanta h^-1^ for standing and speaking (see Figure 1).

**Figure 3.**
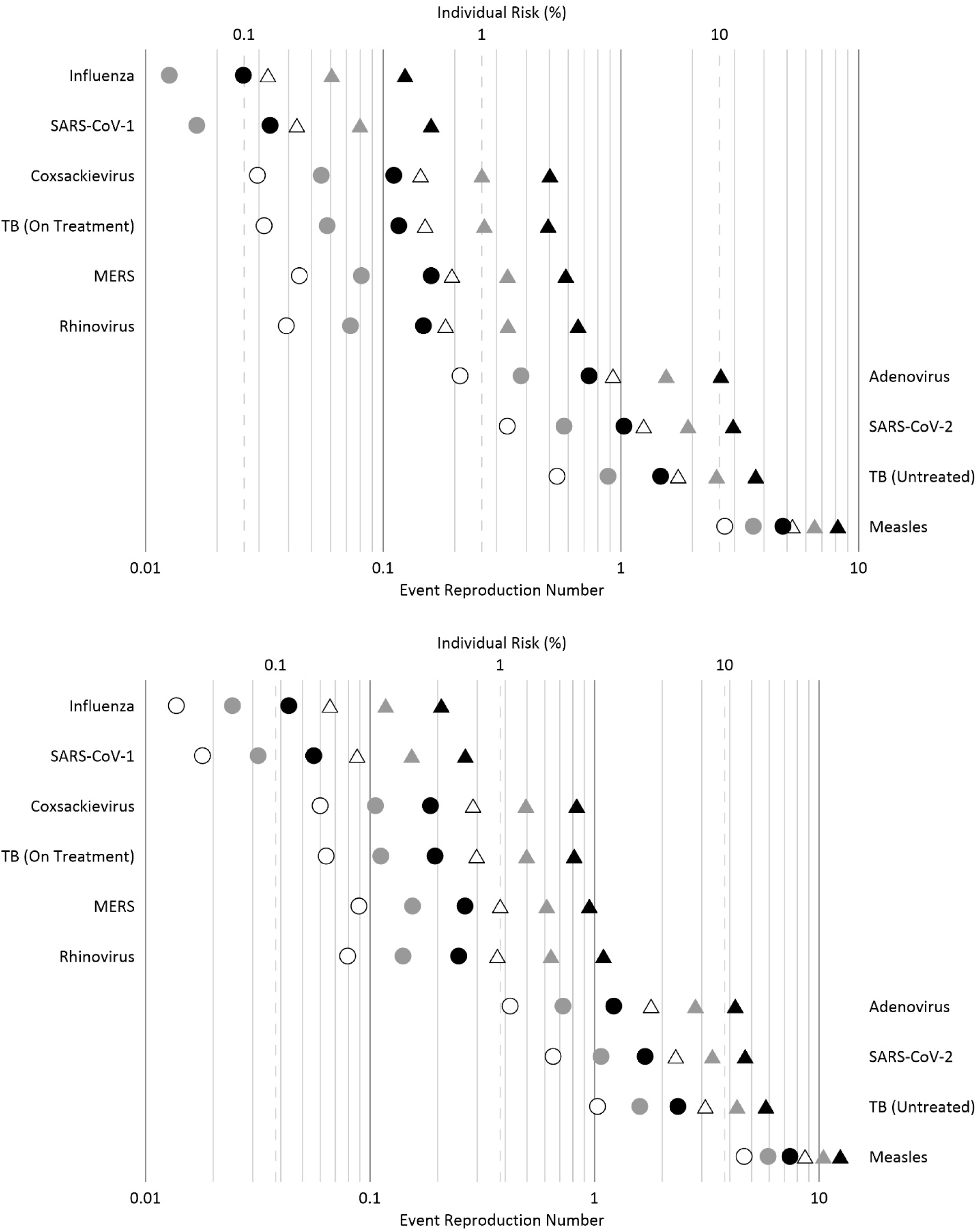
Individual risk (%) and event reproduction numbers (R_event_) for the classroom (3a) and barracks (3b) modeling scenarios. Circles depict results for resting, oral breathing and triangles depict results for standing, speaking. White, gray, and black symbol fill corresponds to the high, medium, and low ventilation rates, respectively.

## 4.0 Conclusions

The ER_q_ estimates we produced for a range of respiratory viruses and for TB are in good agreement with those back calculated from experimental studies and superspreading events in literature. The predictive estimation approach advances methods of prospective risk assessment for airborne transmission of disease. Our analysis should be interpreted as a proof of concept and is limited by the paucity of data on viral load and infectious dose by natural inhalation in the real world. Our calculations suggest measles virus to be the most contagious of those evaluated, but the median estimates for SARS-CoV-2, adenovirus, and untreated, active TB are within the same order of magnitude. The ER_q_ estimates for SARS-CoV-2 and untreated, active TB are quite similar, consistent with their remarkably common settings of contagion. Our risk modeling scenarios for a classroom and barracks show that even a high ventilation rate of 14 L s^-1^ p^-1^ will likely fail to prevent the epidemic spread of adenovirus, TB (untreated), SARS-CoV-2 and measles, indicating that additional mitigation measures such as advanced ventilation design, air disinfection, and reducing the density of susceptibles are necessary. Conversely, this ventilation rate is likely to prevent sustained airborne transmission of rhinovirus, SARS-CoV-1, MERS, coxsackievirus, TB (on treatment) and seasonal influenza.

## Data Availability

N/A

## Supplemental Material

### 1.0 Viral load and infectious dose

Input parameters for the predictive estimation approach for each virus and for tuberculosis (TB) and associated references are summarized in the following sections. We note that in the studies we reviewed the authors often reported infectious dose as being administered via aerosol, or small-particle aerosol. As such, the term aerosol appears in the below summary, and for practical purposes is synonymous with *droplet nuclei* in the context of these studies.

### 1.1 Coronaviruses: SARS-CoV-1, SARS-CoV-2, MERS

Sputum viral load data are available for all three emergent coronaviruses. For SARS-CoV-1, we used an average viral load of 6.1 log_10_ RNA copies mL^-1^ reported by Drosten et al. [1], and for MERS we used the average value of 6.7 log_10_ RNA copies mL^-1^ reported by Corman et al. [2]. For SARS-CoV-2, we calculated an average sputum viral load of 5.6 log_10_ RNA copies mL^-1^ based on the average of values reported by [3-6]. For SARS-CoV-1 and SARS-CoV-2, the standard deviations of the viral load distributions were not explicitly reported in the studies; hence, we calculated the standard deviations such that the 99.9^th^ percentile value of a lognormal distribution (or roughly three standard deviations above the mean) approximately equaled the maximum measured value for SARS-CoV-1, and the average of the maximums for SARS-CoV-2. For MERS, we used the approximate average standard deviation from Corman et al. [2].

For the infectious dose of SARS-CoV-1, we used values of 360 RNA copies PFU^-1^ [7] and 407 PFU quanta^-1^ [8]. For SARS-CoV-2 and MERS, we used estimates of 1.4 x 10^−3^ quanta RNA copies^-1^ and 2.3 x 10^−6^ quanta RNA copies^-1^, respectively, based on the thermodynamic equilibrium dose-response model developed by Gale [9]. In this paper the authors are improving the predictive estimation approach reported in previous works [10, 11] by updating input parameters available in the literature and specific for SARS-CoV-2 such as volume, infectivity and viral load.

### 1.2 Measles and influenza virus

Viral load data for measles in the respiratory tract are limited, and we did not identify a single study where sputum was evaluated. We therefore used the median viral load of 3.5 log_10_ TCID_50_ mL^-1^ in nose swab samples reported in Laksono et al. [12]. The viral load distribution observed by Seto et al. [13] showed greater variation, consistent with the observed pattern of individuals with high viral load capable of infecting large numbers of susceptibles in a shared airspace, or so-called super-spreaders. Hence, we used an approximate log_10_ viral load standard deviation of 1.6, obtained retrospectively from estimates of the number of RNA copies per tube from cycle threshold (C_t_) in the real-time reverse transcription (rRT) polymerase chain reaction (PCR) data set, for which the estimated mean log_10_ viral load was 5.2 log_10_ RNA copies per tube [13].

Data regarding the infectious dose of measles virus are similarly sparse. Considering the highly infectious nature of measles virus, we assumed a conversion factor of 1.0 quanta TCID_50_ ^-1^ based on the minimum infectious dose found for cynomolgus monkeys [14].

For influenza viruses, we used the sputum viral load data reported in Hirose et al. [15] to obtain an average value of 6.7 log_10_ RNA copies mL^-1^ with a log_10_ standard deviation of 0.84. A majority of patients evaluated in the study were diagnosed with the A/H3N2 strain. A conversion factor of 7.1 x 10^−6^ quanta RNA copies^-1^ was selected based on Bueno de Mesquita et al. [16]. This infectious dose is significantly higher than the small-particle aerosol HID_50_ of 0.6-3.0 TCID_50_ reported by Alford et al. [17] using mouthpiece inoculation, reflecting the important distinction between the natural infectious dose by breathing in the real world versus controlled, direct dosage experiments. One should also consider that the typical susceptibility to seasonally circulating influenza virus would be lower than to a novel virus, which may result in a higher dose requirement to initiate infection.

### 1.3 Rhinovirus, adenovirus and coxsackievirus

The viral load of rhinovirus in sputum has been evaluated in studies [18] related to patients with exacerbations of chronic obstructive pulmonary disease (COPD). Since these patients likely have particularly high viral load, they may not be representative of the general population susceptible to rhinovirus. As a result, we used viral load data from nasal wash samples collected during the human transmission trials completed by Meschievitz et al. [19] and Dick et al. [20]. We calculated the average of the geometric mean viral load values reported for each trial as 3.6 log_10_ TCID_50_ mL^-1^ with a log_10_ standard deviation of 0.83. These estimates are consistent with Douglas et al. [21], which studied shedding patterns in rhinovirus-infected volunteers with an average maximum viral titer in nasal wash of 3.2 log_10_ TCID_50_ mL^-1^ with a log_10_ standard deviation of 1.0.

Couch et al. [22] reported the HID_50_ for aerosol inoculation with rhinovirus NIH 1734 as 0.68 TCID_50_. However, the same study notes that an inhaled dose of 2.0 TCID_50_ initially failed to infect any volunteers. Bischoff [23] reported the 100% infectious dose for rhinovirus type 39 via the respiratory tract to be 560 PFUs, or approximately 800 TCID_50_. Therefore, it is likely the HID_63_ representing one quantum falls in-between 2.0 and 800 TCID_50_. Assuming an inhaled dose of 2.0 TCID_50_ is associated with a 10% probability of infection in a Poisson dose-response model, the conversion factor becomes 0.053 quanta TCID_50_^-1^, which we used in the predictive approach.

Airborne transmission of adenovirus and coxsackievirus was studied extensively by the US Army Biological Laboratories based in Fort Detrick, Maryland during the 1960s and early 1970s. Viral load samples were collected from volunteers to support experimental transmission trials and aerosol sampling studies; however, sputum samples were rarely collected. For adenovirus, we used an average viral load of 3.2 log_10_ TCID_50_ mL^-1^ based on throat wash samples reported in Artenstein et al. [24], with a log_10_ standard deviation of 0.95. For coxsackievirus, we used an average viral load of 3.4 log_10_ TCID_50_ mL^-1^ based on nasal secretion samples reported in Buckland et al. [25], with a log_10_ standard deviation of 1.1. We note our average value for adenovirus may be significantly underestimated, as the adenovirus type 4 viral load in sputum reported for one infected subject studied in Couch et al. [26] reached a titer of 6.7 log_10_ TCID_50_ mL^-1^ six days after inoculation, three orders of magnitude higher than the maximum throat swab titer of 3.7 log_10_ TCID_50_ mL^-1^.

Couch et al. [26] reported the HID_50_ for adenovirus type 4 from 1.5 um aerosol inoculation to be 0.5 log_10_ TCID_50_, representing approximately 6 virions. For this reason, Knight [27] stated that adenovirus type 4 in aerosol “approaches the ultimate in infectivity.” As 60% of volunteers administered a reported dose of 1-2 TCID_50_ adenovirus type 4 via aerosol developed infection, we assumed a conversion factor of 0.50 quanta TCID_50_ ^-1^ for the predictive approach. Couch et al. [28] reported the HID_50_ for coxsackievirus A_21_ from 0.3-2.5 um aerosol inoculation to be 28 log_10_ TCID_50_, resulting in a conversion factor of approximately 0.025 quanta TCID_50_ ^-1^ for the predictive approach.

### 1.4 Tuberculosis

Sabiti et al. [29] analyzed the bacillary load of TB in sputum samples from 178 patients at 4 sites in Southeast Africa. Samples were collected weekly, before and during treatment, with a mean estimated bacillary load at baseline of 5.5 log_10_ CFU mL^-1^ declining to approximately 3.0 log_10_ CFU mL^-1^ after 4 weeks of treatment and approximately 1.7 log_10_ CFU mL^-1^ after 8 weeks of treatment [29]. The reported log_10_ standard deviation of estimated sputum bacillary load ranged from 1.2 to 1.4 for different sampling times. To apply the predictive approach to TB, we used both the baseline bacillary load to represent untreated cases, and the approximate average bacillary load after 2 weeks of treatment, or 4.0 log_10_ CFU mL^-1^ [29]. Log_10_ standard deviations of 1.3 and 1.4 were used for untreated and treated TB predictive estimates, respectively.

As summarized by Escombe et al. [30], the infectious dose of TB for humans is unknown, but for fully virulent strains just one droplet nucleus can establish infection and disease in guinea pigs. Bronchoscopic inoculation of 17 cynomolgus macaques with ∼∼25 CFU of TB resulted in evidence of infection in all monkeys with a range of outcomes such as latency mimicking that in humans [31]. Using the low-dose mouse model, Saini et al. [32] reported an equivalency of 2.8 CFU per infectious quantum. However, intentionally efficient laboratory inoculation does not represent natural transmission in the real world. TB infection is an incredibly complex, heterogeneous process, involving aspects such as exogenous reinfection, adaptive immunity, immune exhaustion from repeat exposures, and a characteristic latency, meaning the infectious dose must vary person to person and can be defined and interpreted differently [33]. Indeed, approximately two billion people may have latent TB infection, and there is a greater risk of progressive TB disease with a greater dose of inhaled infectious aerosol [34].

To apply the predictive approach, we used a conversion factor of 2.0 x 10^−3^ quanta CFU^-1^ based on the dose administered to rhesus macaques in Mehra et al. [35]. This reference was selected because the animals were presented with TB using a head-only aerosol method, which is more reflective of natural human-to-human transmission than methods used in other animal studies, and because a 500 CFU aerosol dose resulted in active TB in one of six exposed animals, with the remaining five experiencing latent infection [35]. Therefore, 500 CFU very approximately represents a threshold dose to initiate active infection for rhesus macaques in this experiment.

## 2.0 Evaluation of ERq for coxsackievirus

We used the Gammaitoni and Nucci [36] equation coupled with a Poisson dose-response model to develop an original estimate of ER_q_ for coxsackievirus A_21_ based on the cross-infection experiment in a military barracks documented in Couch et al. [37]. ER_q_ was back calculated using equations 1-3 as follows:

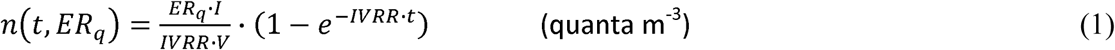

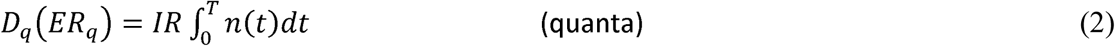

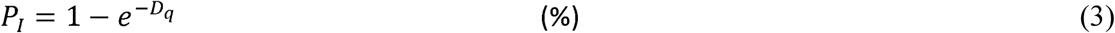

Where *n* represents the quanta concentration in air at time t, ER_q_ is the quanta emission rate (quanta h^-1^), I is the number of infectious subjects, *V* is the volume of the indoor environment considered, *IVRR* (h^-1^) represents the infectious virus removal rate in the space investigated, D_q_ is the dose of quanta inhaled by susceptible persons, T is the total time of the exposure, and P_I_ is the probability of infection of a susceptible person, also commonly termed the attack rate or contact rate. The infectious virus removal rate is the sum of the air exchange rate (AER) via ventilation, the particle deposition on surfaces (*k*, e.g. via gravitational settling), and the viral inactivation (λ).

The barracks consisted of a single room measuring 7.3 x 21 x 3.0 meters (m) separated at its midpoint by a 1.4 m deep double wire netting barrier to prevent close contact between occupants on either side. Ventilation with outside air was minimized by keeping all windows and doors shut with radiant heat only for temperature control. Therefore, we assumed an air exchange rate typical of naturally ventilated buildings of 0.5 hr^-1^. Indoor temperature and relative humidity ranged between 16 and 27 °C and relative humidity between 38 and 68 %. Large floor fans produced well-mixed air throughout the barracks as demonstrated by fluorescein-containing aerosol testing [37].

Twenty volunteers were assigned to one side of the barracks (side A), and 19 to the other side (side B). Ten volunteers on side A only were inoculated with 71 TCID_50_ of coxsackievirus A_21_ to initiate the airborne transmission experiment, with the remaining volunteers receiving a sham inoculation. Evaluation for illness was performed twice daily after inoculation by physicians for the 26-day duration of the study. Extensive measures were taken to separate the residents of side A and side B outside of the barracks, including having them work in completely different parts of the base, ride in different vehicles, and wear arm bands for identification.

All 29 exposed volunteers receiving the sham inoculation became infected with A_21_ by the end of the study. Six susceptible volunteers began to shed virus on day six, constituting a 21% attack rate for this generation. Five of the six initial cross-infected volunteers were on side B and exposed only through the shared air of the barracks. Coxsackievirus was recovered in air samples collected on side B at a concentration of at least 10 TCID_50_ m^-3^ three days prior to the simultaneous detection of virus in five exposed volunteers on side B, consistent with the incubation period of the virus. Hence, we assume the 21% attack rate was produced by an exposure approximately 12 hours in length representing the time spent in one 24-hour day together in the barracks.

Based on the above assumptions and using equations 1-3 we calculated a total emission rate of 27 quanta h^-1^ of coxsackievirus A_21_ produced together by the ten infectious subjects, or 2.7 quanta h^-1^ per person. If the air exchange rate were doubled to 1.0 h^-1^, this emission rate would need to increase to 3.6 quanta h^-1^ per person to generate the 21% attack rate, and if instead the exposure time interval were doubled to 24 hours, this emission rate would decrease to 1.3 quanta h^-1^ per person. Therefore, we consider a range of 1.3 to 3.6 quanta h^-1^ to be a reasonable ER_q_ approximation for coxsackievirus A_21_ based on this experiment.

## 3.0 Calculation of the Individual Risk and Event Reproduction Number for Classroom and Barracks Scenario

With all other parameters held constant, the probability of infection calculated in equation (3) assumes different values based on ER_q_. To evaluate the individual risk (R) of infection of an exposed person for a given exposure scenario, we need to quantify the probability of infection as a function of ER_q_ (P_I_(ER_q_)) and the probability of occurrence of each ER_q_ value (P_ERq_) which can be defined by the probability density function (pdf_ERq_) derived from the predictive estimation approach. Since the probability of infection (P_I_(ER_q_)) and the probability of occurrence P_ERq_ are independent events, R for a given ER_q_, R(ER_q_), can be evaluated as the product of the two terms:

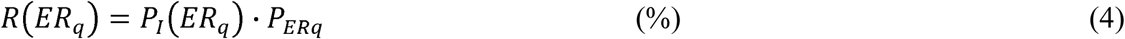

where P_I_(ER_q_) is the conditional probability of the infection, given a certain ER_q_, and P_ERq_ represents the relative frequency of the specific ER_q_ value. The individual risk (R) of an exposed person is then calculated by integrating the pdf_R_ for all possible ER_q_ values, i.e. summing up the R(ER_q_) values calculated in eq. (5):

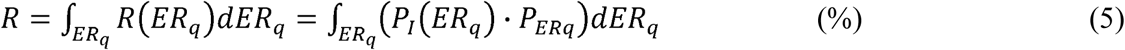

The individual risk R also represents the ratio between the number of new infections and the number of exposed susceptible individuals (S) for a given exposure scenario and taking into account all possible ER_q_ values from an emission distribution for the infectious subject under investigation. For a single exposure event involving a single infectious individual, the number of new infections can be termed the event reproduction number (R_event_), and is calculated as the product of R and S. To facilitate the comparison of contagiousness, we used equations 1-5 to calculate R and R_event_ for each virus and TB for a scenario representing the typical classroom in suburban USA studied by Wells [38, 39], and for the military barracks studied by Couch et al. [37].

For each pathogen and setting we calculated R and R_event_ using the lognormal ER_q_ distributions for two expiratory activity levels, resting and oral breathing and standing and speaking. All susceptible occupants were modeled to be at rest with an inhalation rate of 0.49 m^3^ h^-1^. We modeled the classroom with a room volume of 170 m^3^ and an occupancy of 26 susceptible students and one infectious student for 5.5 hours. For the military barracks, we used the same assumptions as outlined in Section 2.0, but with only one infectious occupant and 38 susceptible occupants for a 12-hour exposure period. A constant deposition rate was used for all pathogens, evaluated as the ratio between the settling velocity of super-micrometric particles (roughly 1.0 × 10^−4^ m s^-1^ [40]) and the height of the emission source (1.5 m). For the inactivation rate in aerosol, we used a value of 0.63 h^-1^ for all pathogens based on the measurements for SARS-CoV-2 reported by van Doremalen et al. [41]. While this inactivation rate likely varies significantly between pathogens and based on environmental conditions such as relative humidity, the comprehensive evaluation of aerosol stability for all viruses and TB is beyond the scope of this paper. Therefore, we used the uniform value to avoid biasing results for any one particular pathogen.

Three air exchange rates (AER) were evaluated for the classroom and barracks scenarios corresponding to ventilation rates of 2.3 liters per second per person (L s^-1^ p^-1^), 6.6 L s^-1^ p^-1^, and 14 L s^-1^ p^-1^. Based on the room volumes these ventilation rates are equivalent to 1.3, 3.8 and 8.0 air changes per hour for the classroom, and 0.70, 2.0 and 4.3 air changes per hour for the barracks, respectively. The results of the modeling scenarios for R_event_ are presented in Supplemental Tables 1 and 2, below. Results are presented graphically in Figure 3 of the main paper. To obtain the individual risk (R), divide R_event_ by the susceptible occupancy of 26 for the classroom and 38 for the barracks.

**Supplemental Table 1.** Event reproduction numbers (R_event_) for the classroom modeled under high ventilation (AER = 8.0 h^-1^), medium ventilation (AER = 3.8 h^-1^), and low ventilation (1.3 h^-1^) conditions. The average R_event_ and individual risk (R) for all classroom scenarios are also provided for reference.

| Expiratory Activity | Resting, Oral Breathing |  |  | Standing, Speaking |  |  | Average |  |
| --- | --- | --- | --- | --- | --- | --- | --- | --- |
| Air Exchange Rate | 8.0 h<sup>-1</sup> | 3.8 h<sup>-1</sup> | 1.3 h<sup>-1</sup> | 8.0 h<sup>-1</sup> | 3.8 h<sup>-1</sup> | 1.3 h<sup>-1</sup> | R<sub>event</sub> | R |
| Influenza | 0.01 | 0.01 | 0.03 | 0.03 | 0.06 | 0.12 | 0.04 | 0.17% |
| SARS-CoV-1 | 0.01 | 0.02 | 0.03 | 0.04 | 0.08 | 0.16 | 0.06 | 0.22% |
| Coxsackievirus | 0.03 | 0.05 | 0.11 | 0.14 | 0.26 | 0.50 | 0.18 | 0.71% |
| TB (On Treatment) | 0.03 | 0.06 | 0.12 | 0.15 | 0.27 | 0.49 | 0.19 | 0.72% |
| MERS | 0.04 | 0.08 | 0.16 | 0.19 | 0.33 | 0.59 | 0.23 | 0.90% |
| Rhinovirus | 0.04 | 0.07 | 0.15 | 0.18 | 0.33 | 0.66 | 0.24 | 0.92% |
| Adenovirus | 0.21 | 0.38 | 0.73 | 0.93 | 1.55 | 2.64 | 1.07 | 4.13% |
| SARS-CoV-2 | 0.33 | 0.58 | 1.03 | 1.25 | 1.91 | 2.96 | 1.34 | 5.17% |
| TB (Untreated) | 0.54 | 0.88 | 1.47 | 1.74 | 2.53 | 3.70 | 1.81 | 6.96% |
| Measles | 2.73 | 3.60 | 4.81 | 5.26 | 6.53 | 8.17 | 5.19 | 19.9% |

**Supplemental Table 2.** Event reproduction numbers (R_event_) for the barracks modeled under high ventilation (AER = 4.3 h^-1^), medium ventilation (AER = 2.0 h^-1^), and low ventilation (0.7 h^-1^) conditions. The average R_event_ and individual risk (R) for all barracks scenarios are also provided for reference.

| Expiratory Activity<br>Air Exchange Rate | Resting, Oral Breathing |  |  | Standing, Speaking |  |  | Average |  |
| --- | --- | --- | --- | --- | --- | --- | --- | --- |
| | 4.3 $\text{h}^{-1}$ | 2.0 $\text{h}^{-1}$ | 0.7 $\text{h}^{-1}$ | 4.3 $\text{h}^{-1}$ | 2.0 $\text{h}^{-1}$ | 0.7 $\text{h}^{-1}$ | $R_{event}$ | R |
| Influenza | 0.01 | 0.02 | 0.04 | 0.07 | 0.12 | 0.21 | 0.08 | 0.21% |
| SARS-CoV-1 | 0.02 | 0.03 | 0.06 | 0.09 | 0.15 | 0.27 | 0.10 | 0.27% |
| TB (On Treatment) | 0.06 | 0.11 | 0.19 | 0.29 | 0.49 | 0.83 | 0.33 | 0.86% |
| Coxsackievirus | 0.06 | 0.11 | 0.19 | 0.30 | 0.50 | 0.81 | 0.33 | 0.87% |
| MERS | 0.09 | 0.15 | 0.26 | 0.38 | 0.61 | 0.95 | 0.41 | 1.07% |
| Rhinovirus | 0.10 | 0.17 | 0.28 | 0.42 | 0.66 | 1.10 | 0.45 | 1.19% |
| Adenovirus | 0.42 | 0.72 | 1.22 | 1.79 | 2.81 | 4.24 | 1.87 | 4.91% |
| SARS-CoV-2 | 0.65 | 1.07 | 1.68 | 2.30 | 3.34 | 4.69 | 2.29 | 6.02% |
| TB (Untreated) | 1.03 | 1.59 | 2.35 | 3.12 | 4.31 | 5.79 | 3.03 | 7.98% |
| Measles | 4.64 | 5.92 | 7.41 | 8.65 | 10.5 | 12.4 | 8.25 | 21.7% |

## References

[1] Balachandar S, Zaleski S, Soldati A, Ahmadi G, Bourouiba L. Host-to-host airborne transmission as a multiphase flow problem for science-based social distance guidelines. International Journal of Multiphase Flow 2020; 132:103439.

[2] Buonanno G, Stabile L, Morawska L. Estimation of airborne viral emission: quanta emission rate of SARS-CoV-2 for infection risk assessment. Environment International 2020; 141:105794.

[3] Wells WF. Airborne Contagion and Air Hygiene. Cambridge, Mass.: Harvard University Press; 1955:423.

[4] Nardell EA. Wells revisited: infectious particles vs. quanta of mycobacterium tuberculosis infection–don’t get them confused. Mycobact Dis 2016; 6(5):1000231.

[5] Bueno de Mesquita PJ, Noakes CJ, Milton DK. Quantitative aerobiologic analysis of an influenza human challenge-transmission trial. Indoor Air 2020; 00:1– 10.

[6] Buonanno G, Morawska L, Stabile L. Quantitative assessment of the risk of airborne transmission of SARS-CoV-2 infection: Prospective and retrospective applications. Environment International 2020; 145:106112.

[7] Stadnytskyi V, Bax CE, Bax A, Anfinrud P. The airborne lifetime of small speech droplets and their potential importance in SARS-CoV-2 transmission. Proc Natl Acad Sci USA 2020; 117(22):11875–11877.

[8] Adams WC. Measurement of Breathing Rate and Volume in Routinely Performed Daily Activities. Final Report. Human Performance Laboratory, Physical Education Department, University of California, Davis. Prepared for the California Air Resources Board, 1993; Contract No. A033–205.

[9] Kulkarni H, Smith CM, Lee DDH, Hirst RA, Easton AJ, O’Callaghan C. Evidence of respiratory syncytial virus spread by aerosol: time to revisit infection control strategies? Am J Respir Crit Care Med 2016; 194:306–16.

[10] Gale P. Thermodynamic equilibrium dose-response models for MERS-CoV infection reveal a potential protective role of human lung mucus but not for SARS-CoV-2. Microb Risk Anal 2020; 16:100140.

[11] Wells WF. Air disinfection in day schools. American Journal of Public Health 1943; 33:1436–1443.

[12] Couch RB, Douglas Jr. RG, Lindgren KM, Gerone PJ, Knight V. Airborne transmission of respiratory infection with coxsackievirus A type 21. American Journal of Epidemiology 1970; 91(1):78–86.

[13] Tupper P, Boury H, Yerlanov M, Colijn C. Event-specific interventions to minimize COVID-19 transmission. Proceedings of the National Academy of Sciences 2020; 202019324.

[14] Zhu S, Jenkins S, Addo K, et al. Ventilation and laboratory confirmed acute respiratory infection (ARI) rates in college residence halls in College Park, Maryland. Environment International 2020; 137:105537.

[15] ASHRAE, ANSI/ASHRAE Standard 62.1-2019. Ventilation for acceptable indoor air quality. Atlanta, GA: American Society of Heating, Refrigerating, and Air-Conditioning Engineers, Inc.; 2019: 92.

[16] Liao CM, Chang CF, Liang HM. A probabilistic transmission dynamic model to assess indoor airborne infection risks. Risk Analysis 2005; 25:1097–107.

[17] Bazant M, Bush JWM. Beyond six feet: a guideline to limit indoor airborne transmission of COVID-19. medRxiv 2020.08.26.20182824 [Preprint]. November 2, 2020 [cited 2020 Dec 23]. Available from: https://doi.org/10.1101/2020.08.26.20182824

[18] Prentiss M, Chu A, Berggren KK. Superspreading events without superspreaders: using high attack rate events to estimate N_0_ for airborne transmission of COVID-19. medRxiv 2020.10.21.20216895 [Preprint]. October 23, 2020 [cited 2020 Dec 23]. Available from: https://doi.org/10.1101/2020.10.21.20216895

[19] Kriegel M, Bucholz U, Gastmeier P, et al. Predicted infection risk for aerosol transmission of SARS-CoV-2. medRxiv 2020.10.08.20209106 [Preprint]. November 5, 2020 [cited 2020 Dec 23]. Available from: https://doi.org/10.1101/2020.10.08.20209106

[20] Miller SL, Nazaroff WW, Jimenez JL, et al. Transmission of SARS-CoV-2 by inhalation of respiratory aerosol in the Skagit Valley Chorale superspreading event. Indoor Air, 2020; 00:1– 10.

[21] Riley RL, Mills CC, O’Grady F, et al. Infectiousness of air from a tuberculosis ward. Ultraviolet irradiation of infected air: comparative infectiousness of different patients. Am Rev Respir Dis 1962; 85:511–525.

[22] Riley EC. The Role of Ventilation in the Spread of Measles in an Elementary School. In: Annals of the New York Academy of Sciences. Airborne Contagion. Vol 353. USA: The New York Academy of Sciences; 1980:25–34.

[23] Azimi P, Keshavarz Z, Cedeno Laurent JG et al. Estimating the nationwide transmission risk of measles in US schools and impacts of vaccination and supplemental infection control strategies. BMC Infect Dis 2020; 20(1):497.

[24] Remington PL, Hall WN, Davis IH, Herald A, Gunn RA. Airborne transmission of measles in a physician’s office. JAMA 1985; 253(11):1574–1577.

[25] Zhou J, Wei J, Choy KT, et al. Defining the sizes of airborne particles that mediate influenza transmission in ferrets. Proc Natl Acad Sci USA 2018; 115(10):E2386–E2392.

[26] Rudnick SN, Milton DK. Risk of indoor airborne infection transmission estimated from carbon dioxide concentration. Indoor Air 2003; 13:237–45.

[27] Moser MR, Bender TR, Margolis HS, Noble GR, Kendal AP, Ritter DG. An outbreak of influenza aboard a commercial airliner. Am J Epidemiol 1979; 110(1):1–6.

[28] Dick EC, Jennings LC, Mink KA, Wartgow CD, Inhorn SL. Aerosol transmission of rhinovirus colds. J Infect Dis 1987; 156(3):442–8.

[29] Langmuir AD. Changing Concepts of Airborne Infection of Acute Contagious Diseases: A Reconsideration of Classic Epidemiologic Theories. In: Annals of the New York Academy of Sciences. Airborne Contagion. Vol 353. USA: The New York Academy of Sciences; 1980:35–44.

[30] Bischoff WE, Swett K, Leng I, Peters TR. Exposure to influenza virus aerosols during routine patient care. The Journal of Infectious Diseases 2013; 207(7):1037–1046.

[31] Russell KL, Broderic MP, Franklin SE, et al. Transmission dynamics and prospective environmental sampling of adenovirus in a military recruit setting. J Infect Dis 2006; 194:877– 85.

[32] Echavarria M, Kolavic SA, Cersovsky S, et al. Detection of adenoviruses (AdV) in culture-negative environmental samples by PCR during an AdV-associated respiratory disease outbreak. J Clin Microbiol 2000; 38(8):2982–2984.

[33] Guo Z, Tong L, Xu S, Li B, Wang Z, et al. Epidemiological analysis of an outbreak of an adenovirus type 7 infection in a boot camp in China. PLOS ONE 2020; 15(6):e0232948.

[34] Nardell EA, Keegan J, Cheney SA, Etkind SC. Airborne infection. Theoretical limits of protection achievable by building ventilation. Am Rev Respir Dis 1991; 144:302–6.

[35] Ko G, Thompson KM, Nardell EA. Estimation of tuberculosis risk on a commercial airliner. Risk Analysis 2004; 24:379–88.

[36] Riley RL, Mills CC, Myka W, et al. Aerial dissemination of pulmonary tuberculosis a two-year study of contagion in a tuberculosis ward. American Journal of Epidemiology 1959; 70(2):185–196.

[37] Escombe AR, Moore DAJ, Gilman RH, et al. The infectiousness of tuberculosis patients coinfected with HIV. PLOS Medicine 2008; 5(9):e188.

[38] Dharmadhikari AS, Mphahlele M, Stoltz A, et al. Surgical face masks worn by patients with multidrug-resistant tuberculosis: impact on infectivity of air on a hospital ward. American Journal of Respiratory and Critical Care Medicine 2012; 185(10): 1104–1109.

## References

[1] Drosten C, Chiu LL, Panning M, et al. Evaluation of advanced reverse transcription-PCR assays and an alternative PCR target region for detection of severe acute respiratory syndrome-associated coronavirus. J Clin Microbiol 2004; 42(5):2043–7.

[2] Corman VM, Albarrak AM, Omrani AS, et al. Viral shedding and antibody response in 37 patients With Middle East Respiratory Syndrome Coronavirus infection. Clin Infect Dis 2016; 62(4):477–483.

[3] Fajnzylber, J., Regan, J., Coxen, K. et al. SARS-CoV-2 viral load is associated with increased disease severity and mortality. Nat Commun 2020; 11:5493.

[4] Pan Y, Zhang D, Yang P, Poon LLM, Wang Q. Viral load of SARS-CoV-2 in clinical samples. Lancet Infect Dis 2020; 20(4):411–412.

[5] Wölfel, R., Corman, V.M., Guggemos, W. et al. Virological assessment of hospitalized patients with COVID-2019. Nature 2020; 581:465–469.

[6] To KK, Tsang OT, Leung WS, et al. Temporal profiles of viral load in posterior oropharyngeal saliva samples and serum antibody responses during infection by SARS-CoV-2: an observational cohort study. The Lancet. Infectious Diseases 2020; 20(5):565–574.

[7] Vicenzi E, Canducci F, Pinna D, et al. Coronaviridae and SARS-associated coronavirus strain HSR1. Emerg Infect Dis 2004; 10(3):413–418.

[8] Watanabe T, Bartrand TA, Weir MH, Omura T, Haas CN. Development of a dose-response model for SARS coronavirus. Risk Anal 2010; 30:1129–1138.

[9] Gale P. Thermodynamic equilibrium dose-response models for MERS-CoV infection reveal a potential protective role of human lung mucus but not for SARS-CoV-2. Microb Risk Anal 2020; 16:100140.

[10] Buonanno G, Stabile L, Morawska L. Estimation of airborne viral emission: quanta emission rate of SARS-CoV-2 for infection risk assessment. Environment International 2020; 141:105794.

[11] Buonanno G, Morawska L, Stabile L. Quantitative assessment of the risk of airborne transmission of SARS-CoV-2 infection: Prospective and retrospective applications. Environment International 2020; 145:106112.

[12] Laksono BM, de Vries RD, Verburgh RJ, et al. Studies into the mechanism of measles-associated immune suppression during a measles outbreak in the Netherlands. Nat Commun 2018; 9:4944.

[13] Seto J, Ikeda T, Tanaka S, et al. Detection of modified measles and super spreader using a real-time reverse transcription PCR in the largest measles outbreak, Yamagata, Japan, 2017 in its elimination era. Epidemiology and Infection 2018; 146:1707–1713.

[14] van Binnendijk RS, van der Heijden RW, van Amerongen G, UytdeHaag FG, Osterhaus AD. Viral replication and development of specific immunity in macaques after infection with different measles virus strains. J Infect Dis 1994; 170(2):443–448.

[15] Hirose R, Daidoji T, Naito Y, et al. Long-term detection of seasonal influenza RNA in faeces and intestine. Clin Microbiol Infect 2016; 22(9):813.e1-813.e7.

[16] Bueno de Mesquita PJ, Noakes CJ, Milton DK. Quantitative aerobiologic analysis of an influenza human challenge-transmission trial. Indoor Air 2020; 00:1– 10.

[17] Alford RH, Kasel JA, Gerone PJ, Knight V. Human influenza resulting from aerosol inhalation. Proc Soc Exp Biol Med 1966; 122:800–804.

[18] Mallia P, Message SD, Gielen V, et al. Experimental rhinovirus infection as a human model of chronic obstructive pulmonary disease exacerbation. Am J Respir Crit Care Med 2011; 183(6):734–42.

[19] Meschievitz C, Schultz S, Dick E. A model for obtaining predictable natural transmission of rhinoviruses in human volunteers. J Infect Dis 1984; 150:195

[20] Dick EC, Jennings LC, Mink KA, Wartgow CD, Inhorn SL. Aerosol transmission of rhinovirus colds. J Infect Dis 1987; 156(3):442–8.

[21] Douglas R, Cate T, Gerone P, Couch R. Quantitative rhinovirus shedding patterns in volunteers. -Amer Rev Respiratory Dis 1966; 94:159–167.

[22] Couch RB, Cate TR, Douglas RG Jr, Gerone PJ, Knight V. Effect of route of inoculation on experimental respiratory viral disease in volunteers and evidence for airborne transmission. Bacteriol Rev 1966; 30(3):517–29.

[23] Bischoff WE. Transmission route of rhinovirus type 39 in a monodispersed airborne aerosol. Infect Control Hosp Epidemiol 2010; 31(8):857–859.

[24] Artenstein MS, Miller WS, Lamson TH, Brandt BL. Large-volume air sampling for Meningococci and Adenoviruses. Am J Epidemiol 1968; 87(3):567–77.

[25] Buckland FE, Bynoe ML, Tyrrell DA. Experiments on the spread of colds. II. Studies in volunteers with coxsackievirus A21. J Hyg (Lond) 1965; 63(3):327–343.

[26] Couch RB, Knight V, Douglas RG Jr, Black SH, Hamory BH. The minimal infectious dose of adenovirus type 4; the case for natural transmission by viral aerosol. Trans Am Clin Climatol Assoc 1969; 80:205–211.

[27] Knight V. Viruses as Agents of Airborne Contagion. In: Annals of the New York Academy of Sciences. Airborne Contagion. Vol 353. New York, USA: The New York Academy of Sciences; 1980:147–156.

[28] Couch RB, Cate TR, Gerone PJ, et al. Production if illness with a small-particle aerosol of coxsackie A21. J Clin Invest 1965; 44(4):535–542.

[29] Sabiiti W, Azam K, Farmer ECW, et al. Tuberculosis bacillary load, an early marker of disease severity: the utility of tuberculosis Molecular Bacterial Load Assay. Thorax 2020; 75(7):606–608.

[30] Escombe AR, Moore DAJ, Gilman RH, et al. The infectiousness of tuberculosis patients coinfected with HIV. PLOS Medicine 2008; 5(9):e188.

[31] Capuano SV 3rd, Croix DA, Pawar S, et al. Experimental Mycobacterium tuberculosis infection of cynomolgus macaques closely resembles the various manifestations of human M. tuberculosis infection. Infect Immun 2003; 71(10):5831–5844.

[32] Saini D, Hopkins GW, Seay SA, et al. Ultra-low dose of Mycobacterium tuberculosis aerosol creates partial infection in mice. Tuberculosis (Edinb) 2012; 92(2):160–5.

[33] Nardell EA. Wells revisited: infectious particles vs. quanta of mycobacterium tuberculosis infection–don’t get them confused. Mycobact Dis 2016; 6(5):1000231.

[34] Fennelly KP, Jones-López EC. Quantity and quality of inhaled dose predicts immunopathology in Tuberculosis [published correction appears in Front Immunol. 2015;6:511]. Front Immunol 2015; 6:313.

[35] Mehra S, Golden NA, Dutta NK, et al. Reactivation of latent tuberculosis in rhesus macaques by coinfection with simian immunodeficiency virus. J Med Primatol 2011; 40(4):233–243.

[36] Gammaitoni L, Nucci MC. Using a mathematical model to evaluate the efficacy of TB control measures. Emerging Infectious Diseases 1997; 3(3):335–342.

[37] Couch RB, Douglas Jr. RG, Lindgren KM, Gerone PJ, Knight V. Airborne transmission of respiratory infection with coxsackievirus A type 21. American Journal of Epidemiology 1970; 91(1):78–86.

[38] Wells WF. Air disinfection in day schools. American Journal of Public Health 1943; 33:1436–1443.

[39] Wells WF. Airborne Contagion and Air Hygiene. Cambridge, Mass.: Harvard University Press; 1955:423.

[40] Chatoutsidou SE, Lazaridis M. Assessment of the impact of particulate dry deposition on soiling of indoor cultural heritage objects found in churches and museums/libraries. Journal of Cultural Heritage 2019; 39:221–228.

[41] van Doremalen N, Bushmaker T, Morris DH, et al. Aerosol and surface stability of SARS-CoV-2 as compared with SARS-CoV-1. N Engl J Med 2020; 382(16):1564–1567.

